# CASP4/11 contributes to pulmonary inflammation and disease exacerbation in COVID-19

**DOI:** 10.1101/2022.06.29.22277067

**Authors:** Tamara S. Rodrigues, Camila C.S. Caetano, Keyla S.G. de Sá, Leticia Almeida, Amanda Becerra, Augusto V. Gonçalves, Leticia de Sousa Lopes, Samuel Oliveira, Danielle P.A. Mascarenhas, Sabrina S. Batah, Bruna M. Silva, Giovanni F. Gomes, Ricardo Castro, Ronaldo B. Martins, Jonathan Avila, Fabiani G. Frantz, Thiago M. Cunha, Eurico Arruda, Fernando Q Cunha, Helder Nakaya, Larissa D. Cunha, Alexandre T Fabro, Paulo Louzada-Junior, Renê D.R. de Oliveira, Dario S. Zamboni

## Abstract

Infection with SARS-CoV-2 induces COVID-19, an inflammatory disease that is usually self-limited, but depending on patient conditions may culminate with critical illness and patient death. The virus triggers activation of intracellular sensors, such as the NLRP3 inflammasome, which promotes inflammation and aggravates the disease. Thus, identification of host components associated with NLRP3 inflammasome is key for understanding the physiopathology of the disease. Here, we reported that SARS-CoV-2 induces upregulation and activation of human Caspase-4/CASP4 (mouse Caspase-11/CASP11) and this process contributes to inflammasome activation in response to SARS-CoV-2. CASP4 was expressed in lung autopsy of lethal cases of COVID-19 and CASP4 expression correlates with expression of inflammasome components and inflammatory mediators such as *CASP1*, *IL1B*, *IL18* and *IL6*. In vivo infections performed in transgenic hACE2 humanized mouse, deficient or sufficient for *Casp11*, indicate that hACE2 *Casp11^−/−^* mice were protected from disease development, with reduced body weight loss, reduced temperature variation, increased pulmonary parenchymal area, reduced clinical score of the disease and reduced mortality. Collectively, our data establishes that CASP4/11 contributes to disease pathology and contributes for future immunomodulatory therapeutic interventions to COVID-19.

## Introduction

Human infections with SARS-CoV-2 emerged in the end of 2019 and culminated with the massive COVID-19 pandemic, leading to more than half billion cases and 6 million deaths worldwide. The disease manifests as mild to asymptomatic in most patients, but ineffective innate immune responses early after infection can lead to inflammatory complications that may evolve to death (Blanco-Melo et al., 2020; Madden and Diamond, 2022; Merad et al., 2022; Wiersinga et al., 2020). Complications in severe to critical cases of COVID-19 are often immuno-mediated, rather than caused by viral replication itself.

Hence, although antiviral therapies are already available for patient treatment, novel immunomodulatory approaches are required to effectively treat patients with severe disease. Thus, understanding the mechanisms involved in exacerbated inflammation in critical cases of COVID-19 is essential to develop more effective treatments. One of the immune signatures identified in severe cases of COVID-19 is the activation of intracellular proteins that promotes inflammation, called inflammasomes (Lucas et al., 2020). According to these findings, it was demonstrated that SARS-CoV-2 triggers NLRP3 inflammasome upon infection of human monocytes and the inflammasome is highly active in mild to severe COVID-19 patients (Junqueira et al., 2022; Rodrigues et al., 2021; Sefik et al., 2022).

The NLRP3 inflammasome is activated in the cytoplasm of mammalian cells in response to pathogenic microbes or in response to cell stress or membrane perturbations. Once activated, NLRP3 receptor oligomerizes and recruits the adaptor molecule ASC, which polymerizes and recruits caspase-1 that is proteolytically activated and promotes cleavage of many inflammatory substrates including pro-IL-1β, pro-IL-18 and Gasdermin-D (Broz and Dixit, 2016; He et al., 2016; Shi et al., 2017). As consequence, inflammasome-activated cells release inflammatory cytokines and undergo an inflammatory form of cell death called pyroptosis. Mechanistically, the canonical NLRP3 inflammasome activation occurs in response to molecules that directly induce pore formation or damage membranes, allowing efflux of K^+^-mediated NLRP3 activation (Broz and Dixit, 2016; He et al., 2016; Shi et al., 2017). In contrast, activation of human Caspase-4/CASP4 (mouse Caspase-11/CASP11) leads to the non-canonical activation of NLRP3, a process that requires an intracellular pore-forming molecule to promote the release of K^+^ and enable NLRP3 activation (Baker et al., 2015; Ruhl and Broz, 2015; Schmid-Burgk et al., 2015). Molecules that trigger CASP4/11 activation in the cytoplasm of mammalian cells include bacterial lipopolysaccharide, parasites lipophosphoglycan and endogenous molecules such as oxidized phospholipids (de Carvalho et al., 2019; Hagar et al., 2013; Kayagaki et al., 2013; Zanoni et al., 2016). Despite the importance of pyroptosis in antiviral immunity (reviewed in (Kuriakose and Kanneganti, 2019)), the role of CASP4/11 in viral infections remains poorly understood. For SARS-CoV-2 infection, it was demonstrated that NLRP3 inflammasome is activated in response to infection, but the mechanisms underlying this activation and the molecules that participate in this process are still unclear. Here, we demonstrated that CASP4/11 participates in the inflammasome activation in response to SARS-CoV-2 and promotes exacerbation of the disease in mouse models of infection. Our results effectively account for understanding the physiopathology of the COVID-19 and establish CASP4 as a potential target for immune-mediated therapies to treat patients in moderate and critical conditions.

## Results

### Caspase-4 is upregulated in COVID-19 patients and activated during SARS-CoV-2 infection

To investigate whether *CASP4* is upregulated in patients with COVID-19 and how this correlates with disease severity, we assessed RNA sequencing data from nasopharyngeal swab of 10 healthy donor controls and COVID-19 patients, including 58 that tested positive for the virus (non-hospitalized), 17 that were hospitalized but did not required ICU and 8 that were admitted to intensive care unit (ICU) (Ng et al., 2021). The expression of *CASP4* was significantly higher in COVID-19 patients when compared to controls, regardless of the degree of disease severity (**Fig. 1A, B**). Inflammasome-associated genes such as *CASP1*, *IL1B* and IL-18 family members (*IL18*, *IL18R1*, *IL18RAP*) were also upregulated and positively correlated with *CASP4* in COVID-19 patients (**Fig. 1B-D**). CASP4 is transcriptionally regulated during infections and proteolytically cleavage during activation (Rathinam et al., 2019). Thus, we tested CASP4 cleavage in PBMCs obtained from fresh blood samples of COVID-19 patients collected during hospitalization (all moderate to severe cases of COVID-19). We detected CASP4 cleavage in PBMC obtained from COVID-19 patients (in 7 out of 13 patients tested) (**Fig. 1E**). To further assess CASP4 activation in response to SARS-CoV-2 infection, we measured caspase-4 activation in primary human monocytes infected in vitro. We found that infection with viable SARS-CoV-2, but not with UV irradiated virus induced caspase-4 activation as measured by the substrate Ac-WEHD-FC (**Fig. 1F**). The NLRP3 inhibitor MCC950, which effectively inhibits SARS-CoV-2-induced NLRP3 activation (Rodrigues et al., 2021) was used as control and indicate a non-relevant participation of NLRP3/caspase-1 activity in this assay. Altogether, these data indicated that *CASP4* gene expression is upregulated in COVID-19 patients and SARS-CoV-2 infection triggers CASP4 activation in monocytes infected *in vitro* and in PBMCs of COVID-19 patients.

**Figure 1.**
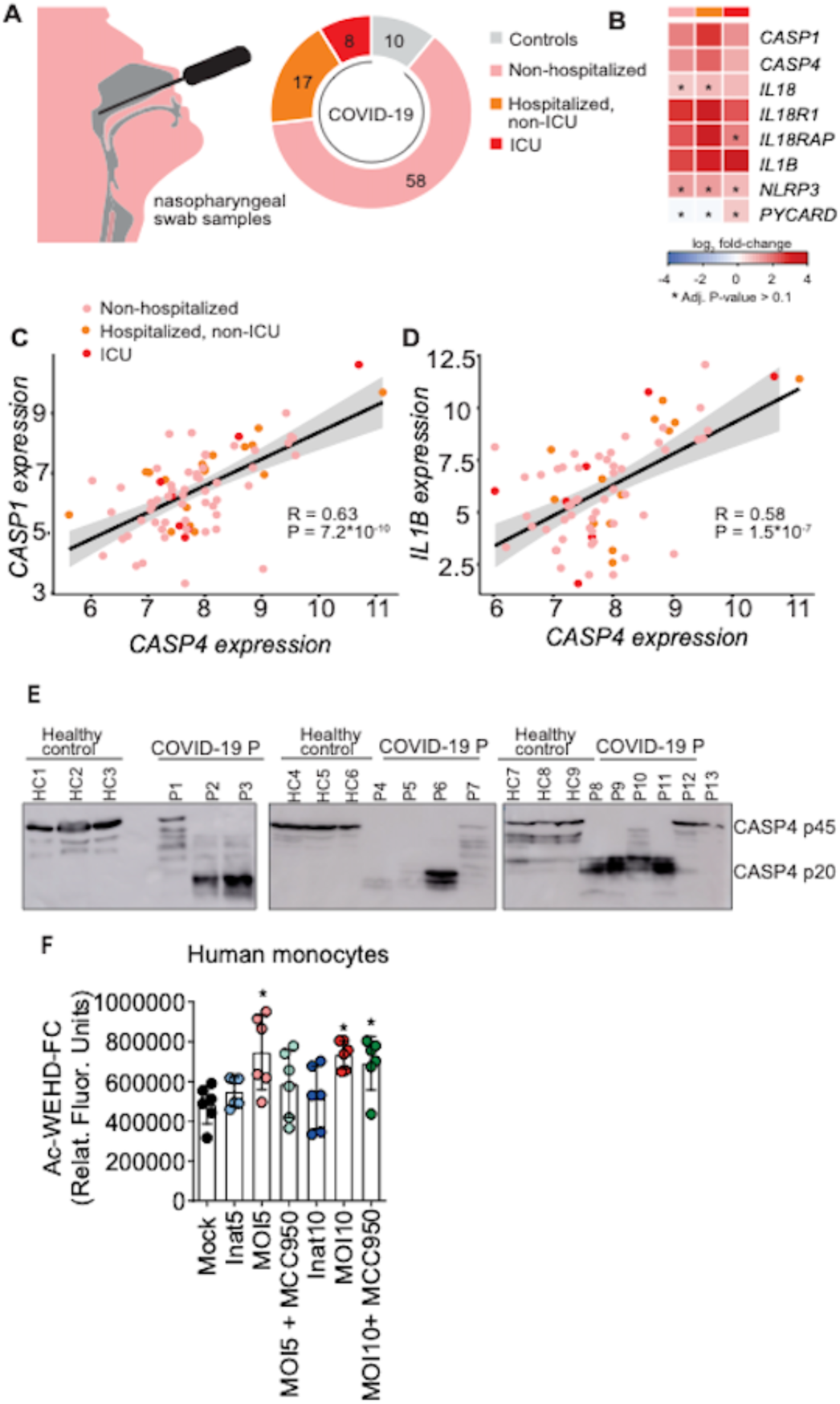
Caspase-4 is upregulated COVID-19 patients and activated during SARS-CoV-2 infection. **(A-D)** Caspase-4 (*CASP4*) expression was evaluated in nasopharyngeal swab samples from controls and from COVID-19 patients divided in three groups: Non-hospitalized; Hospitalized, non-ICU; and ICU **(A)**. *CASP4* expression was correlated with other inflammasome-related genes **(B)**. *CASP4* correlation with *CASP1* **(C)** and *IL1B* **(D)**, indicating Non-hospitalized; Hospitalized, non-ICU; and ICU. **(E)** Peripheral Blood Mononuclear Cells (PBMCs) were isolated from fresh blood of Healthy Controls (HC 1-9, n=9) or COVID-19 P (P1-13, n=13). Caspase-4 cleavage was assessed by western blotting analysis of cell lysate. **(F)** Human CD14+ monocytes were primed with PAM3Cys (300ng/mL) for 4 hours and infected with SARS-CoV-2 at a multiplicity of infection (MOI) of 1 and 5 for 24 h. Mock was used as a negative infection control. Virus was inactivated by UV irradiation (U.V. Inat.) at each specific MOI. The NLRP3 inhibitor MCC950 (10 μM) was added 1 h after infection and maintained. At 24 hours post infection, caspase-4 activation was performed using the caspase-4 fluorescent substrate Ac-WEHD-AFC. *, *P* < 0.05 compared to MOCK and U.V. Inat. MOI5 as determined by Student’s t-test. Box shows average ± SD of the values.

### Caspase-4 is upregulated in the lungs of lethal cases of COVID-19 and correlates with inflammatory mediators and inflammasome components

Next, we assessed the expression of CASP4 and inflammatory mediators in lung autopsies of lethal cases of COVID-19. By performing RT-PCR we found that CASP4 is upregulated in the lungs of COVID-19 patients (n=28) as compared to control patients (benign area of lung adenocarcinoma, n=4) (**Fig. 2A**). It is worth noticing that many patients did not show *CASP4* expression, indicating that CASP4 expression and activation is variable among COVID-19 patients. All 28 patients shown in this figure were blood culture negative, suggesting that bacterial or fungi co-infections may not contribute to increased CASP4 expression observed in patients. Additional analysis including samples from COVID-19 patients that tested positive for fungi or bacteria during hospitalization does not support a significant contribution of co-infections for the increased *CASP4* expression (**Supplementary Fig. 1**). Interestingly, the RT-PCR analyses of gene expression in culture-negative COVID-19 patient showed a positive correlation between *CASP4* and several inflammatory genes (**Fig. 2B**). Among those, we found a significant positive correlation with *CASP1*, *IL1B*, *IL18*, *IL6*, *TNFA* (**Fig. 2C-G**). By performing sequential immunoperoxidase labeling and erasing method in the pulmonary tissues, we confirmed the expression of CASP4, CASP1 and SARS-CoV-2 Spike proteins in the lungs of fatal COVID-19 patients (**Fig. 2H**).

**Figure 2.**
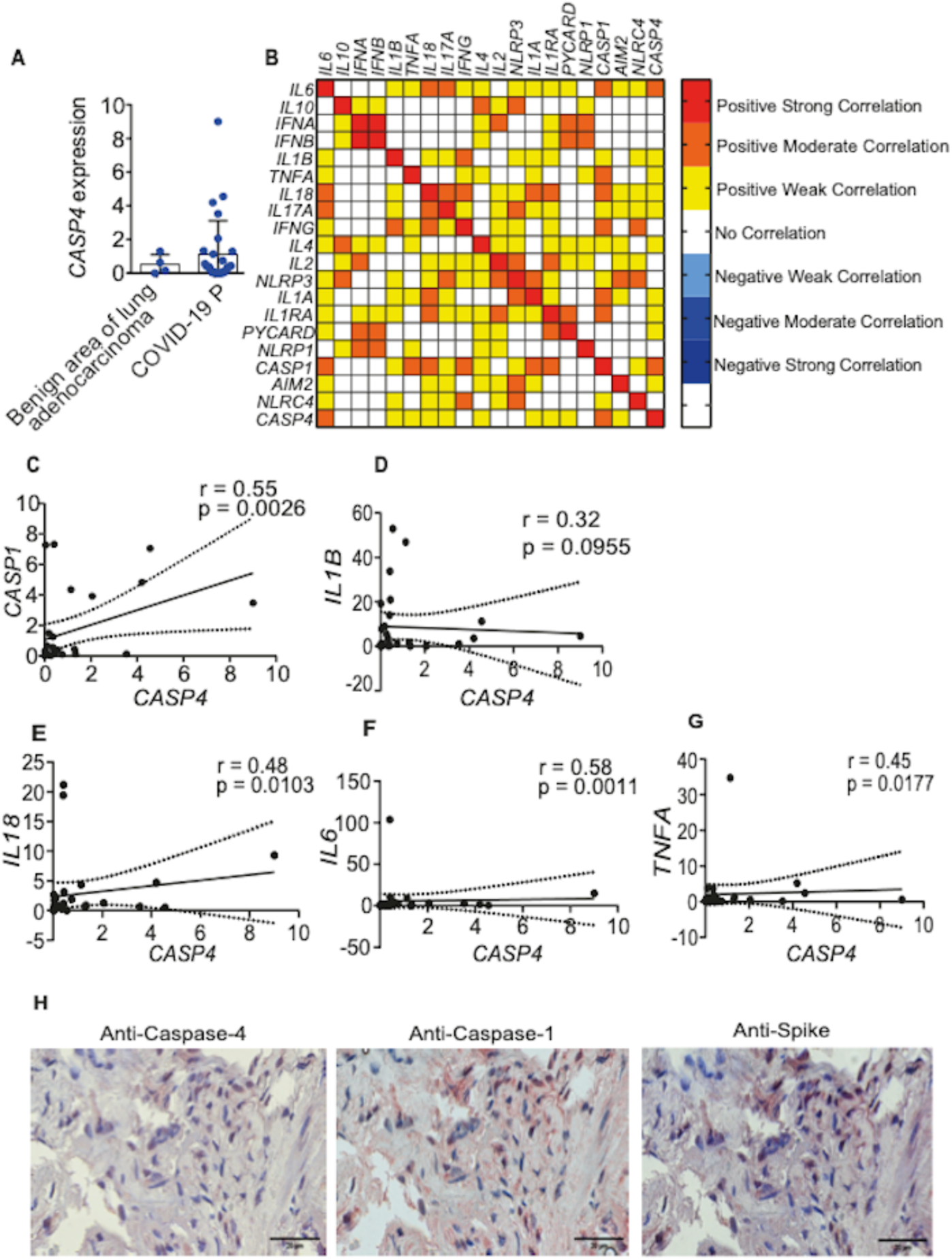
Caspase-4 is upregulated in the lungs of lethal cases of COVID-19 and correlates with inflammatory mediators and inflammasome components. Lung autopsies were obtained from 28 fatal cases of COVID-19 that tested negative for co-infections (blood culture) during hospitalization. Controls includes samples from 4 patients that deceased due to lung adenocarcinoma (benign area of lungs). **(A-G)** The expression of caspase-4 mRNA (*CASP4*) and other inflammatory genes were assessed in pulmonary autopsies of COVID-19 patients by RT-PCR. **(B)** Correlation matrix of CASP4 gene expression and other inflammatory genes indicating positive strong correlation (red), positive moderate correlation (orange) and positive weak correlation (yellow). **(C-G)** Correlations of *CASP4* with *CASP1* (C), IL1B **(D)**, IL-18 **(E)**, IL-6 **(F**) and TNF **(G)**. **(H**) Multiplex staining by sequential immunohistochemistry staining illustrates CASP4 expression (anti-Caspase-4). CASP1 expression (anti-Caspase-1) and SARS-CoV-2 Spike protein (anti-Spike) in pulmonary tissue of one COVID-19 patient. Scale bar 20 μm.

### Caspase-11 is important for inflammasome activation in macrophages infected with SARS-CoV-2

CASP11 activation in mouse cells triggers the non-canonical activation of the NLRP3 inflammasome (Kayagaki et al., 2011). Thus, we investigated the effect of CASP11 in the activation of NLRP3 induced by SARS-CoV-2 infection. Initially, we tested if SARS-CoV-2 infection induces upregulation of CASP11 in mouse bone marrow-derived macrophages (BMDMs) and found that viable SARS-CoV-2 infection, but not UV-irradiated virus, induced CASP11 expression (**Fig. 3A**). Infection of *Nlrp3^−/−^* BMDMs revealed that NLRP3 was not required for CASP11 upregulation (**Fig. 3A**).

**Figure 3.**
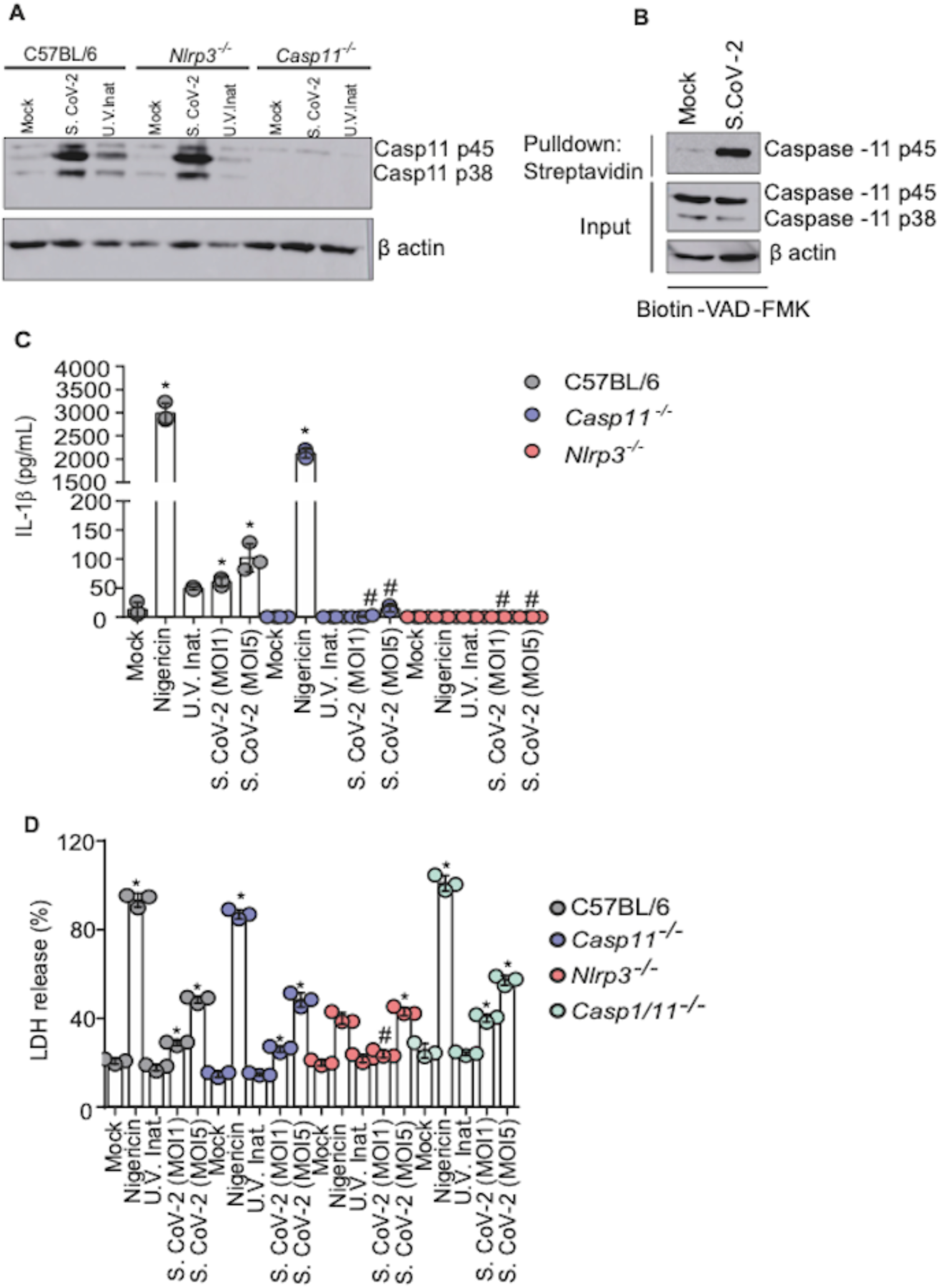
Caspase-11 is important for inflammasome activation in macrophages infected with SARS-CoV-2. **(A)** Bone marrow derived macrophages (BMDMs) were obtained from C57BL/6, *Nlrp3^−/−^* and *Casp11^−/−^* mice. BMDMs were infected with SARS-CoV-2 at a multiplicity of infection (MOI) of 1 for 8h. Western blotting analysis of cell lysate was performed to evaluate caspase-11 expression after infection. **(B)** C57BL/6 BMDMs were primed with PAM3Cys (300 ng/mL) for 4 hours and infected with SARS-CoV-2 at a MOI of 1 for 8h. Pull-down of active caspase-11 was performed by addition of Biotin-VAD-FMK followed by Streptavidin beads. (**C**) BMDMs were obtained from C57BL/6 K18-hACE-2 and mice backcrossed with *Nlrp3^−/−^* and *Casp11^−/−^* mice. Cells were primed with PAM3Cys (300 ng/mL) for 4 hours and infected with SARS-CoV-2 at MOI of 1 for 24 and IL-1β levels in cell-free supernatants was measured by ELISA. **(D)** BMDMs from C57BL/6, *Nlrp3^−/−^, Casp11^−/−^* and *Casp1/11^−/−^* in the K18-hACE-2 background were infected with SARS-CoV-2 at a MOI of 1 for 24h and LDH release was measured in the supernatants. * indicates *P* < 0.05 compared to MOCK in the same genetic background and ^#^ indicates *P* < 0.05 compared to C57BL/6 in the same conditions. Both determined by Student’s t-test. Box shows average ± SD of the values.

To further investigate the activation of CASP11 in response to infection, we performed a streptavidin pull-down assay in Biotin-VAD-FMK treated cells and probed active caspase-11 by western blot (Cunha et al., 2015). Using this assay, we found that SARS-CoV-2 infection triggers CASP11 activation in BMDMs (**Fig. 3B**). To investigate the importance of CASP11 for the NLRP3 inflammasome activation in response to SARS-CoV-2, we infected BMDMs and measured IL-1β production as a readout for inflammasome activation. We found that SARS-CoV-2, but not UV-inactivated virus, triggered IL-1β production in C57BL/6 macrophages and this process was impaired in *Casp11^−/−^* macrophages (**Fig. 3C**). The bacterial toxin Nigericin, an activator of canonical NLRP3 inflammasome, was used as a positive control. These data indicates that SARS-CoV-2 triggers activation of the inflammasome in a process dependent on NLRP3 and CASP11. Finally, we tested the effect of NLRP3 and CASP11 for the lytic form of cell death induced by SARS-CoV-2. By measuring LDH release, we found that NLRP3, CASP11 and CASP1/11 are dispensable for cell lysis induced by SARS-CoV-2 (**Fig. 3D**). This data is consistent with previously reported studies (Rodrigues et al., 2021) and indicates that additional pathways (besides inflammasomes) participate in the induction of macrophage death in response to SARS-CoV-2 infection.

### Caspase-11 mediates disease pathology in ACE2-humanized mice infected with SARS-CoV-2

To access the importance of CASP11 in host response to SARS-CoV-2 in vivo, we generated a K18-hACE2 transgenic mice (Zheng et al., 2021a) deficient in *Casp11*. Initially, we tested the effect of CASP11 in viral replication in vivo. Thus, hACE2 *Casp11^+/+^*, *Casp11^+/–^* and *Casp11^−/−^* littermate control mice were infected with SARS-CoV-2 2×10^4^ PFU and the viral load was estimated during infection. We did not detect any effect of CASP11 in viral replication when we measured viral loads by TCID50 or RT-PCR (**Fig. 4A-B**). Next, we evaluated the importance of CASP11 for inflammasome activation in vivo. We assessed NLRP3 and ASC puncta formation, a process that indicates activation of the NLRP3 inflammasome and occurs in response to SARS-CoV-2 infection and in COVID-19 patients (Rodrigues et al., 2021). We confirmed that SARS-CoV-2-infected mice induces puncta in the lungs of hACE2 *Casp11^+/+^* mice and this process is reduced in *Casp11^−/−^* littermate controls (**Fig. 4 C-D**). MOCK-infected mice show non-significant puncta formation. To the best of our knowledge, this is the first data demonstrating activation of NLRP3 inflammasome in response to SARS-CoV-2 in mouse models of COVID-19. Importantly, Caspase-11 is required for efficient inflammasome activation in mouse lungs. **Fig. 4 E-F** illustrate NLRP3 and ASC puncta in the lungs of SARS-CoV-2-infected mice. According to the puncta formation data, we found a reduced IL-1β production in *Casp11^−/−^* compared to *Casp11^+/+^* at 3 days of infection (**Supplementary Fig. 2A**). Additional cytokines were measured, including IFNγ, IL-6, IL-10, IL-12, TNFα and MCP1. Although they were slightly lower in *Casp11^−/−^* mice, they were not statistically significant compared to *Casp11^+/+^* and *Casp11^+/−^* mice (**Supplementary Fig. 2B-G**).

**Figure 4.**
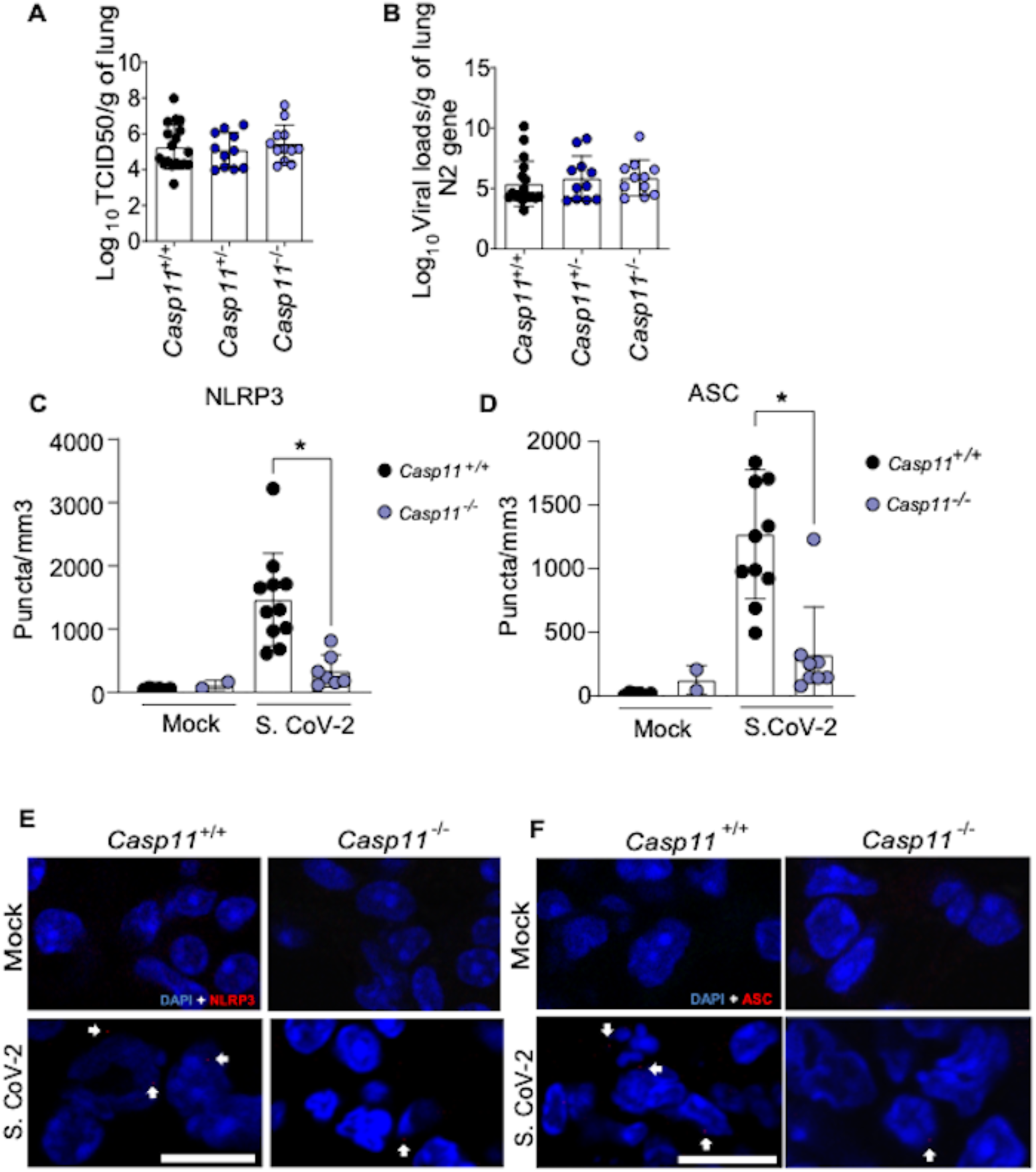
Caspase-11 does not influence viral replication but it is important for inflammasome activation in ACE2-humanized mice infected with SARS-CoV-2. Transgenic K18-hACE2 mice heterozygous for *Casp11* were crossed to generate K18-hACE2 *Casp11^+/+^*, *Casp11^+/–^* and *Casp11^−/−^* littermate control mice. Animals were infected intranasally with 2×10^4^ SARS-CoV-2 per mice for 3 days. Viral load was evaluated by TCID50 **(A)** and by RT-PCR for SARS-CoV-2 N2 gene **(B)**. The percentage of cells containing NLRP3 **(C)** and ASC **(D)** puncta was estimated by multiphoton microscopy of pulmonary tissues. Multiphoton microscopy images of pulmonary tissues stained with anti NLRP3 (**E**) or ASC (**F**) indicates inflammasome puncta (red dots, indicated by white arrows). DAPI stains cell nuclei. Scale bar 10 μm.

Next, we investigated the effect of CASP11 in disease development and found that *Casp11^−/−^* mice were protected from weight loss compared to *Casp11^+/+^* and *Casp11^+/−^* littermate controls (**Fig. 5A**). *Casp11^−/−^* mice were also more resistant to temperature variation (**Fig. 5B**) and show a reduced clinical score during disease development (**Fig. 5C-D**). To further evaluate the pulmonary status of SARS-CoV-2-infected mice, we performed histological analyses of the infected lungs. By assessing H&E-stained sections of the lung tissues, we found reduced cellular infiltrate in *Casp11^−/−^* mice with reduced parenchyma area as compared to *Casp11^+/+^* mice infected for 5 days (**Fig. 5E-F**). Similar results were obtained in histological analyses of mice infected for 3 days (**Supplementary Fig. 3**). Finally, we tested the mice mortality during course development. Initially, we monitored mice for up to 10 days after infection and found that SARS-CoV-2-infected mice started to die on day 6 and *Casp11^−/−^* were significantly more resistant to death as compared to *Casp11^+/+^* and *Casp11^+/−^* littermate controls (**Fig. 5G**). This figure shows a pool of 2 independent experiments with n=23 *Casp11^+/+^*; n=16 *Casp11^+/−^*; n=28 *Casp11^−/−^*. We found that at day 10 a few mice were sick and we decided to perform longer experiments. Following mice survival for up to 16 days after infection, we confirmed that *Casp11^−/−^* were significantly more resistant than *Casp11^+/+^*. Whereas *Casp11^+/+^* died by day 8, *Casp11^+/−^* died by day 14 and about 30% of the *Casp11^−/−^* survived and did not appear sick (**Fig. 5H**). These in vivo data strongly indicate that CASP11 participates in the disease exacerbation and may contribute to the excessive inflammation observed in the lungs of critical cases of COVID-19.

**Figure 5.**
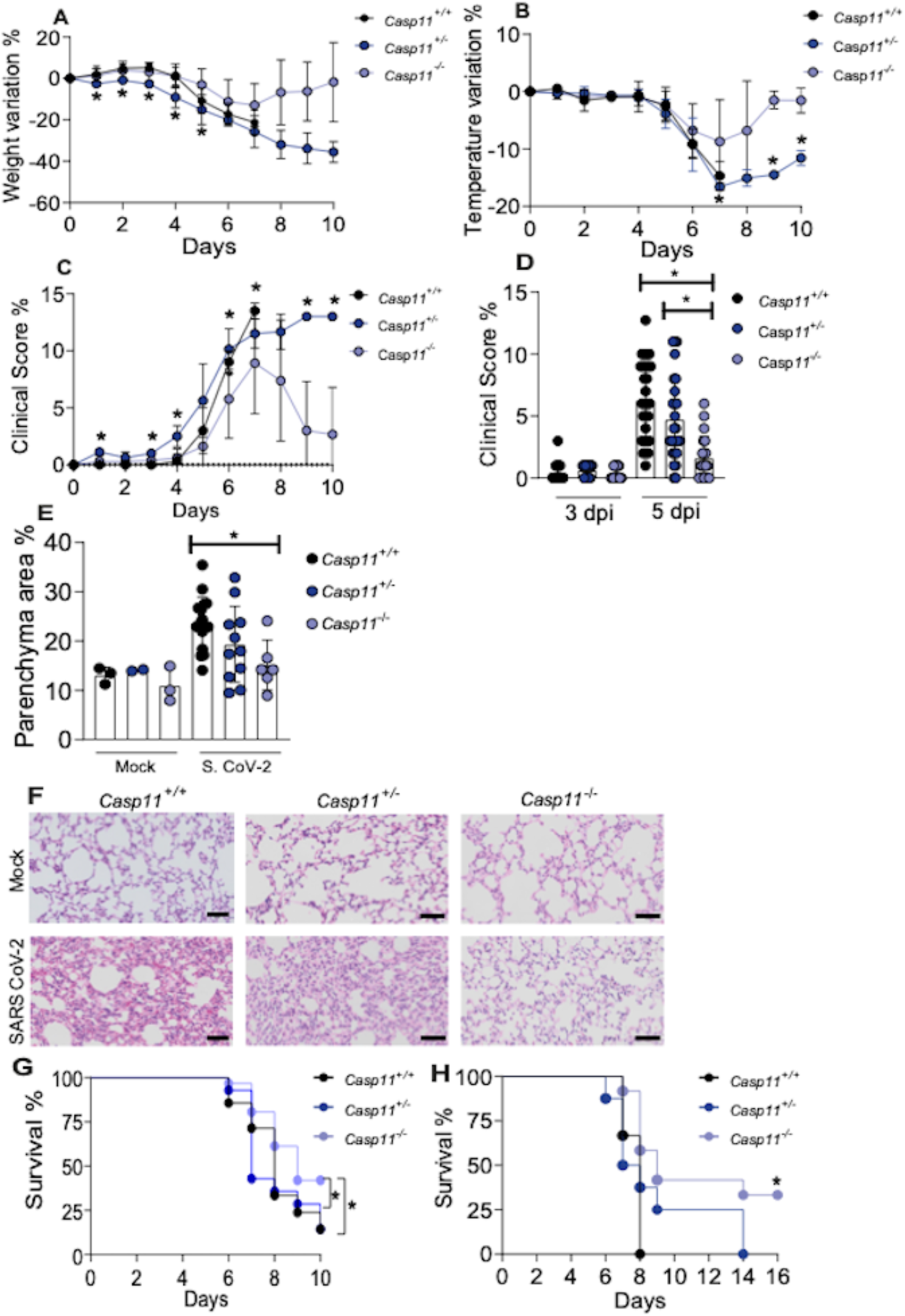
Caspase-11 mediates disease pathology in ACE2-humanized mice infected with SARS-CoV-2. Transgenic K18-hACE2 mice heterozygous for *Casp11* were crossed to generate K18-hACE2 *Casp11^+/+^*, *Casp11^+/–^* and *Casp11^−/−^* littermate control mice. Animals were infected intranasally with 2×10^4^ SARS-CoV-2 per mice for up to 10 days. Body weight variation **(A)**, temperature variation **(B)** and clinical score **(C)** were evaluated from day 0 to day 10. Clinical score of 3 and 5 d post infection of a pool from 3 independent experiments **(D). (E)** The percentage of parenchyma area were estimated after hematoxylin and eosin (H&E) staining of lung sections of K18-hACE2 *Casp11^+/+^*, *Casp11^+/−^* and *Casp11^−/−^* infected for 5 days. Box shows average ± SD of the values. Representative images of H&E-stained lungs of mice infected for 5 days are shown (**F**). Upper panels show Mock-infected mice. Scale bar 50 μm. (**G**) survival curve of K18-hACE2 *Casp11^+/+^* (n=23), *Casp11^+/−^* (n=16) and *Casp11^−/−^* (n=28) infected and followed for 10 days (pool of 2 independent experiments. (**H**) Survival curve of K18-hACE2 *Casp11^+/+^* (n=3), *Casp11^+/−^* (n=8) and *Casp11^−/−^* (n=13) infected and followed for 16 days (* in this figure indicates *P* < 0.05 comparing *Casp11^+/+^* with *Casp11^−/−^*). *, *P* < 0.05, as determined by Man Whitney (for 2 groups) or Kruskal-Wallis (for 3 groups) test (**A-D**) or Kaplan-Meier (**G-H**).

## Discussion

Exacerbated inflammasome activation aggravates COVID-19 by promoting excessive pulmonary inflammation and patient death (Batah et al., 2022; Junqueira et al., 2022; Lucas et al., 2020; Rodrigues et al., 2021; Sefik et al., 2022). As the NLRP3 inflammasome is central in critical patients with COVID-19, immunomodulatory strategies to target NLRP3 may allow important therapeutic possibilities to treat critical cases of COVID-19. Thus, the identification of additional molecules that participate in NLRP3 inflammasome activation augments therapeutic targets available to treat severe and critical patients. Here, we identified CASP4/11 as a key host component involved in disease exacerbation. Importantly, all the patients and human samples included in this study were obtained before July 2020, when only ancestral Wuhan strain of SARS-CoV-2 existed and vaccines were not yet available. Thus, the clinical and biological data obtained from patients were not influenced by different viral strains and the presence of specific T and B cells populations. Our data obtained with human samples and in vitro infections were supported by in vivo experiments performed with ACE2-humanized mice, which demonstrated that CASP11 contributes to disease exacerbation. It is interesting that CASP11 did not interfered with viral replication in pulmonary tissues. Either CASP4/11 has a positive and a negative effect in viral replication and they neutralize each other or CASP4/11 activation does not influence any biological process involved in SARS-CoV-2 replication in tissues. It is worth mentioning that these findings were consistent with a recently published study, where the authors show a role of CASP4/11 in disease aggravation, but not in viral replication (Eltobgy et al., 2022). Importantly, the fact that CASP4/11 ablation does not increase viral replication in tissues provides relevant information to endorse further exploitation of drugs that target CASP4 in COVID-19 patients.

CASP4/11 is not highly expressed in steady state tissues and transcriptional regulation of *CASP4/11* during infections are required to guarantee sufficient amount of protein to be proteolytically activated and exerts effector functions. It is interesting that SARS-CoV-2 triggers a potent *CASP4/11* transcriptional response. It is possible that the reported activity of SARS-CoV-2 Spike or Envelope proteins in TLR2/NF-κB signaling account for this process (Khan et al., 2021; Zheng et al., 2021b). Alternatively, cytokines produced in response to viral infection such as IL-6 and TNFα may trigger NF-κB signaling that in turn promote *CASP4/11* expression. Regardless to the mechanism by which the SARS-CoV-2 triggers transcriptional regulation of CASP4/11, this process was consistently observed in macrophage infections in vitro, in nasopharyngeal samples of COVID-19 patients and in the lungs of fatal cases of COVID-19. It is interesting to note that CASP4 expression was very variable among COVID-19 patients (Figs. 1 and 2) and this does not appear to correlate with co-infections (Supplementary Fig. 1). As the virus itself induces CASP4/11 expression, it is possible that differences in viral loads explain differences in CASP4 expression. Further investigation will be required to associate the early expression of CASP4 and viral loads with the clinical outcome of COVID-19 in patients. Regardless to the possible use of CASP4 as clinical predictor of disease severity, the determination that CASP4 participate in disease exacerbation account to our understanding of COVID-19 and offers an important therapeutic target for future immunomodulatory therapy to treat COVID-19 patients in severe and critical conditions.

## Material and Methods

### Ethical Statements

All experiments performed with human samples and with animals were approved by institutional ethics committee. This includes experiments with human blood samples (Research Ethics Committee Protocol from Hospital das Clinicas de Ribeirão Preto - USP: CAAE, n° 06825018.2.3001.5440) and experiments with Lung autopsies (Research Ethics Committee of FMRP/USP under protocol n° 4.089.567). All mice experiments were conducted according to the guidelines from the institutional ethical committees for animal care from Comissão de Ética em Experimentação Animal da Faculdade de Medicina de Ribeirão Preto, FMRP/USP (Approved protocol number 133/2020).

### RNA sequencing data from nasopharyngeal swab of COVID-19 patients

The gene count table and metadata were provided by authors from GEO (GSE163151). Then, differential expression analysis was performed with the DESeq2 package using the default parameters (Love et al., 2014). COVID-19 patients were subdivided in 3 groups according to their disease severity: Non-hospitalized COVID-19 patients (N = 58); Hospitalized COVID-19 patients but not ICU (N =17); and ICU (N=8). Pearson correlation analyses between CASP4 and CASP1 and between CASP4 and IL1B were performed in R using UQ count normalized data, which was generated using the EDAseq package (Risso et al., 2011).

### Patients’ blood samples and Peripheral blood mononuclear cells isolation

PBMCs were isolated from patients with COVID-19 enrolled in Hospital das Clínicas, Faculdade de Medicina de Ribeirão Preto da Universidade de São Paulo from April 6 to July 2, 2020 and tested positive using RT-PCR (Corman et al., 2020; Lu et al., 2020). For peripheral blood mononuclear cells isolation, whole blood was collected from healthy donors or from COVID-19 patients in tubes containing EDTA (BD Vacutainer CPT; BD Biosciences). The blood was centrifuged at 400 x g for 10 minutes at room temperature. Then, the plasma was discarded and the cell pellet was resuspended in PBS 1X pH 7.4 (GIBCO, BRL). The cells were added to the Ficoll-PaqueTM PLUS gradient column (GE Healthcare Biosciences AB). Next, the gradient was centrifuged at 640 x g for 30 minutes at room temperature to obtain the purified mononuclear fraction, which was carefully collected and transferred to a new tube. Then, the PBMCs were washed and the pellet was resuspended in RPMI for following analysis.

### Western blotting for Caspase-4

1×10^7^ PBMCs from Healthy controls (HC, n=9) or COVID-19 patients (n=11) were lysed in 50uL of RIPA (10mM Tris-HCl (pH 7.4), 1mM EDTA, 150mM NaCl, 1% Nonidet P-40, 1% (w/v) sodium deoxycholate and 0.1% (w/v) SDS) supplemented with 4% protease inhibitor cocktail (Roche). Proteins were separated by 15% SDS–polyacrylamide gel electrophoresis, transferred onto a nitrocellulose membrane and the membranes were incubated overnight, at 4°C under mild agitation with mAb rabbit anti-casp4 (Cell Signaling, 1:1000) antibody diluted 1:1000 in 5% BSA in TBS 1X with 0.01% Tween. The membranes were incubated for 1h with goat anti-rabbit HRP secondary antibody (Sigma-Aldrich, Missouri, USA) and analyzed using ECL™ Prime Western Blotting System (GE Healthcare, Illinois, EUA) and an Amersham Imager 600 (GE Healthcare, Illinois, EUA).

### Lung autopsies from fatal cases of COVID-19

From May to July, 2020, 47 patients diagnosed with SARS-CoV-2 at the Hospital das Clínicas da Faculdade de Medicina de Ribeirão Preto da Universidade de São Paulo, Brazil (Ribeirão Preto, SP, Brazil) underwent minimally invasive autopsy within 1 hour of death. A post-mort surgical lung biopsy was performed with a 3 cm incision on the previous side of the chest between the fourth and fifth ribs. A matching 14-gauge cutting needle (Magnum Needles, Bard) and a biopsy gun (Magnum, Bard) were also used. The patients who died from adenocarcinoma between the years 2012 to 2020 were used as controls and tissue samples of lung necropsies were obtained from the Serviço de Patologia (SERPAT) of the Hospital das Clínicas da Faculdade de Medicina de Ribeirão Preto da Universidade de São Paulo, Brazil. Samples were embedded in paraffin and fixed in formalin (Formalin-Fixed Paraffin-Embedded, FFPE).

### Viral Stock Production

The SARS-CoV-2 used was the Brazil/SPBR-02/2020 strain which was propagated under BSL3 conditions. Before infection, Vero CCL81 cells were washed with PBS1x. Then, cells were infected with the viral inoculum using DMEM with trypsin-TPCK treatment (1µg/μL) at 37°C, 5% CO_2_. When the virus-induced cytopathic effect was observed, the cells were harvested, and centrifuged (10.000 x G). The supernatant was stored at −80°C, and the virus titration was performed on Vero CCL81 cells.

### Purification of human monocytes

The monocytes (CD14+ cells) were purified using positive selection with magnetic nanoparticles (BD). Briefly, PBMCs were labeled with BD IMag™ Anti-human CD14 Magnetic Particles – DM and plated cell were placed over a magnetic field for cell separation as previously described (Rodrigues et al., 2020). The positive fraction, purified CD14 + monocytes, were then cultured in RPMI 1640 (GIBCO, BRL) containing 10% SFB.

### Caspase-4 activation assay

1×10^5^ Purified human monocytes were plated in back/clear bottom 96 well plates, and infected with SARS-COV2 MOI1 and MOI5. After 1 hour of viral adsorption, medium (RPMI 2% FBS without Fenol Red) was added. Cells were incubated for 8h at 37°C in the presence of 5% CO_2_. After that, the caspase-4 substrate Ac-WEHD-AFC was added to the cells at a final concentration of 100 μM and cell were incubated up to 24 hours post infection. Substrate cleavage was monitored by measuring the emission at 505 nm on excitation at 400 nm using the SpectraMax i3 system (Molecular Devices).

### RNA extraction and RT-PCR for inflammatory genes

Total RNA from fresh lung tissue of SARS-CoV-2 patients and respective controls was obtained using Trizol reagent and purification was performed according to the manufacturer’s instructions. The RNA was quantified by spectrophotometry in a NanoDrop 2000c spectrophotometer. The concentration was adjusted to 1 ug/µL, and the RNA was stored at −80 ◦C until reverse transcription. For quantification of inflammatory genes, the total RNA was transcribed into complementary DNA (cDNA) using a High-Capacity cDNA Reverse Transcription kit (without an inhibitor) according to the protocol provided by the manufacturer (Thermo Fisher, Carlsbad, CA, USA).

The real-time PCR was performed in 96-well plates using Sybr Green reagents (Applied Biosystems, Waltham, MA, USA) and a Quant studio real-time PCR system (Applied Biosystems, Foster City, CA, USA) following the protocol recommended by the manufacturer. The Ct values were analyzed by the comparative Ct (ΔΔCt) method and normalized to the endogenous control GAPDH. Fold difference was calculated as 2−ΔΔCt. Primers used for human genes included: *GSDMD*: F:ATGAGGTGCCTCCACAACTTCC and R:CCAGTTCCTTGGAGATGGTCTC; *NLRP3:* F:GGACTGAAGCACCTGTTGTGCA and R:TCCTGAGTCTCCCAAGGCATTC; *IL1A*: F:TGTATGTGACTGCCCAAGATGAAG and R:AGAGGAGGTTGGTCTCACTACC; *IL1RA*: F:ATGGAGGGAAGATGTGCCTGTC and R:GTCCTGCTTTCTGTTCTCGCTC; *NLRC4*: F:AGGTCCCACAACTCGTCAAGCT and R:TGCTCACACGATTTCCCGCCAA; *PYCARD*: F:AGCTCACCGCTAACGTGCTGC and R:GCTTGGCTGCCGACTGAGGAG; *NLRP1*: F:ATTGAGGGCAGGCAGCACAGAT and R:CTCCTTCAGGTTTCTGGTGACC; *CASP1*: F:GCTGAGGTTGACATCACAGGCA and R:TGCTGTCAGAGGTCTTGTGCTC; *AIM2*: F:GCTGCACCAAAAGTCTCTCCTC and R:CTGCTTGCCTTCTTGGGTCTCA; *IL6*: F:AGACAGCCACTCACCTCTTCAG and R:TTCTGCCAGTGCCTCTTTGCTG; *GAPDH*: F:GTCTCCTCTGACTTCAACAGCG and R:ACCACCCTGTTGCTGTAGCCAA; *TNFA*: F:CTCTTCTGCCTGCTGCACTTTG and R:ATGGGCTACAGGCTTGTCACTC; *IL10*: F:TCTCCGAGATGCCTTCAGCAGA and R:TCAGACAAGGCTTGGCAACCCA; *IL1B*: F:CCACAGACCTTCCAGGAGAATG and R:GTGCAGTTCAGTGATCGTACAGG; *IL18*: F:GATAGCCAGCCTAGAGGTATGG and R:CCTTGATGTTATCAGGAGGATTCA; *CASP4*: F:GGGATGAAGGAGCTACTTGAGG and R:CCAAGAATGTGCTGTCAGAGGAC; *IL17A*: F:CGGACTGTGATGGTCAACCTGA and R:GCACTTTGCCTCCCAGATCACA; *IFNG*: F:GAGTGTGGAGACCATCAAGGAAG and R:TGCTTTGCGTTGGACATTCAAGTC; *IFNB1*: F:CTTGGATTCCTACAAAGAAGCAGC and R:TCCTCCTTCTGGAACTGCTGCA; *IFNA1*: F:AGAAGGCTCCAGCCATCTCTGT and R:TGCTGGTAGAGTTCGGTGCAGA; *IL4*: F:CCGTAACAGACATCTTTGCTGCC and R:GAGTGTCCTTCTCATGGTGGCT.

### Sequential immunoperoxidase labeling and erasing

Tissue section in paraffin blocks were tested by immunohistochemistry using antibodies for the detection of Spike (1:500, Abcam, ab272504) and Caspase-4 (1:200, MBL, M029-3). The Sequential Immunoperoxidase Labeling and Erasing (SIMPLE) technique was used to evaluate all markers in the same tissue (Glass et al., 2009). After incubation with primary antibody, slides were incubated with immune peroxidase polymer anti-mouse visualization system (SPD-125, Spring Bioscience, 345 Biogen) and with chromogen-substrate AEC peroxidase system kit (SK-4200, 346 Vector Laboratories, Burlingame, CA). After marking, the slides were scanned in a VS120 Olympus microscope. After high resolution scanning the coverslips were removed in PBS and the slides were dehydrated in an ethanol gradient to 95% ethanol. The slides were incubated in a series of ethanol to erase the AEC marking. Afterwards the slides were rehydrated and the antibodies were removed with an incubation for 2 min in a solution of 0.15 mM 351 KMnO4/0.01 M H_2_SO_4_, followed immediately by a wash in distilled water. Tissues were then remarked.

### Bone marrow derived macrophages

Bone marrow-derived macrophages were obtained as previously described (Marim et al., 2010). Briefly, mice were euthanized and bone marrow cells were isolated from femurs. The cells were cultivated in RPMI 1640 (Gibco, Thermo Fisher Scientific, Massachussetts, USA) supplemented with 20% Fetal Bovine Serum (FBS) (Gibco) and 10% of a conditional media from 3T3 cell, which express mouse M-CSF as a source of macrophage colony stimulation factor, for 7 days, at 37°C, 5% CO2. Cells were detached with cold PBS 1x, resuspended in RPMI 1640 supplemented with 10% FBS (R10) and plated as indicated. Incubation of non-infected and infected cells was done at 37°C, 5% CO2.

### Western blotting and active Caspase-11 pull-down assays

To measure caspase-11 expression by western blot, 1×10^6^ BMDMs were infected with SARS-CoV-2 MOI of 1 for 8h. Then, cells were lysed with 40 μL of RIPA supplemented with 4% protease inhibitor cocktail (Roche). For active caspase11, 1×10^6^ cells were primed with TLR2 agonist PAM-3Cys for 4 hours and replenished with fresh media containing 20 mM biotin-VAD-FMK (Enzo) 15 min before infection. Then, cells were infected with SARS-CoV-2 MOI of 1 for 8 hours. Next, BMDMs were washed twice with PBS and cells were lysed in 100uL of RIPA buffer supplemented with protease inhibitor cocktail (Roche). Cleared lysates were equalized according to total protein content, incubated overnight with streptavidin-agarose beads (Invitrogen) and thoroughly rinsed with RIPA buffer. Bound proteins were eluted by re-suspension in Laemmli sample buffer, boiled for 5min and separated by 15% SDS–polyacrylamide gel electrophoresis. After run, proteins are transferred onto a nitrocellulose membrane and the membranes were incubated overnight, at 4°C under mild agitation with mAb Anti-Casp11 (ABCAM) antibody diluted 1:1000 in 5% non-fat dry milk in TBS 1X with 0.01% Tween. β actin was stained with mouse anti-β actin (Santa Cruz 1:3000) diluted in 5% non-fat dry milk in TBS 1X with 0.01% Tween. The membranes were incubated for 1h with goat anti-rabbit HRP or goat anti-mouse HRP secondary antibody (Sigma-Aldrich, Missouri, USA) and analyzed using ECL™ Prime Western Blotting System (GE Healthcare, Illinois, EUA) and an Amersham Imager 600 (GE Healthcare, Illinois, EUA).

### Evaluation of IL-1β and LDH release in cell culture supernatants

2 x 10^5^ bone marrow derived macrophages were plated on 48-well plates in RPMI 10%FBS and incubated overnight. In the following day, cells were primed with PAM3cys 300ng/mL for 4 hours. The PAM3cys stimulus was then removed and cells were infected with SARS-COV-2 using MOI of 1 and MOI of 5. After 1h of adsorption, RPMI without Phenol Red (3.5 g/L HEPES, 2 g/L NaHCO_3_, 10.4 g/L RPMI without Phenol Red, 1% glutamine, pH 7.2) was added to complete media and cells were incubated for 24h. Supernatant was collected to evaluate IL-1β and LDH release. LDH release was measured using CytoTox 96® Non-Radioactive Cytotoxicity Assay (Promega, Winsconsin, USA) following the manufacturer’s instructions. IL-1β was quantified by ELISA (R&D Systems) in the supernatants of in vitro infected macrophages following manufacturer’s instructions.

### Animals and in vivo infections

Mouse used in this study were in C57BL/6 genetic background and included K18-hACE2 (B6.Cg-Tg(K18-ACE2)2Prlmn/J; Jackson Laboratory strain#:034860) that was crossed with *Casp11^−/−^* for generation of *hACE2^+/–^ Casp11^+/–^*, which were crossed for the generation of the *hACE2^+/–^ Casp11^+/+^*, *hACE2^+/–^ Casp11^+/–^*, and *h-ACE2^+/–^ Casp11^−/−^*. Male or Female mice ranging from eight- to ten-weeks-old were infected in a BSL3 facility at the University of São Paulo, FMRP/USP. All animals were provided food and water ad libitum, at 25°C. Infections experiments were performed with 8 to 10 weeks old mice. The animals were anesthetized with ketamine (50mg/kg) and xylazine (10mg/kg) intraperitoneally and infected intranasally with SARS-CoV-2 2×10^4^ PFU contained in 20 μL of PBS was administered intranasally. From day 0 to day 10, animals were evaluated for weight, temperature and clinical score. The clinical score was measured according to weight loss, mice posture and appearance, activity, eyes opening, responsiveness to stimuli, breathing (Supplementary Table 1). At the indicated time point, mice were euthanized and lungs were collected for following analyses. For mortality curves, mice survival was monitored daily with food and water provided *ad libitum*.

### TCID50 and RT-PCR for SARS-CoV-2 assays in mouse

The virus titration of viral stocks of from infected-mouse lung homogenates were performed using standard limiting dilution to confirm the 50% tissue culture infectious dose (TCID50) of the propagated viral stock (Reed, 1938). Viral quantification by RT-PCR, was performed with primer-probe sets for 2019-nCoV_N2, according to USA-CDC and Charité group protocols (Corman et al., 2020; Lu et al., 2020). The N2 gene was analyzed by one-step real-time RT-PCR using total RNA extracted with Trizol (Invitrogen, CA, EUA) from lung homogenates in order to measure the genome viral load from the infected animals. All real-time PCR assays were performed in QuantStudio™ 3 Real-Time PCR System, 96-well, 0.1 mL (Applied Biosystems). Briefly, RNA extraction was performed by Trizol. 100 ng of RNA was used for genome amplification, adding specifics primers (20 µM), and probe (5 µM), and with TaqPath 1-Step qRT-PCR Master Mix (Applied Biosystems, Foster City, CA, USA), with the following parameters: 25°C for 2 min, 50°C for 15 min, 95°C for 2 min, followed by 45 cycles of 94 °C for 5 s and 60 °C for 30s. N2 primers and probe includes: N2-fwd: 5’-TTA CAA ACA TTG GCC GCA AA-3’; N2-rev: 5’-GCG CGA CAT TCC GAA GAA-3’; N2Probe: 5’-FAM-ACA ATT TGC CCC CAG CGC TTC AG-BHQ1-3’.

### Cytokine quantification in mice lung homogenates, histological evaluation and oxygen saturation, breath rate and heart rate measure in mice

IL-1β levels were evaluated in the lung tissue homogenate from the lungs of SARS-CoV-2 infected mice by ELISA. TNF-α, IL-6, IL-12, IL-10, IFNγ, and MCP-1 were quantified in mice lung tissue homogenates by CBA (CBA Mouse Inflammation Kit, BD) following manufacturer’s instructions. For histological evaluation, mice lung tissue samples were fixed in formalin 10%, paraffin-embedded and standard haematoxylin and eosin (H&E) stained in 3 μm sections. The tissue slides were scanned on a VS120 Olympus microscope with high resolution in order to perform histological analysis of parenchyma area by measuring aerated area percent using imageJ software. The mice oxygen saturation, breath rate and heart rate were measured daily in non-anesthetized animals using the MouseOx Plus, according to manufacturer instructions.

## Statistical Analysis

Statistical significance of * P<0.05 was determined by two-tailed unpaired Student t test for data that reached normal distribution or by the non-parametric Mann–Whitney or Kruskall-Wallis tests for not normally distributed data. Unpaired t test was performed for the in vitro experiments. Kruskall-Wallis with Dunn’s correction was performed for in vivo data with 3 group comparisons while Mann-Whitney was used for the 2 group comparisons. Kaplan-Meier test was performed for survival curves. These statistical procedures and graph plots were performed with GraphPad Prism 8.4.2 software. The normal distribution in samples of lung gene expression was evaluated using the Shapiro–Wilk test; the data were analyzed using non-parametric Kruskal-Wallis, Mann-Whitney test and Spearman correlation.

## Data Availability

All data produced in the present study are available upon reasonable request to the authors.

## Acknowledgments

We would like to thank Maira Nakamura, Debora Perucello, Amanda Zuin and Roberta Sales for technical support.

## Funding sources

Fundação de Amparo à Pesquisa do Estado de Sao Paulo, FAPESP grants 2013/08216-2, 2019/11342-6 and 2020/04964-8. Conselho Nacional de Desenvolvimento Científico e Tecnológico, CNPq grant 303021/2020-9. Coordenação de Aperfeiçoamento de Pessoal de Nível Superior, CAPES grant 88887.507253/2020-00.

## Competing interests

Authors declare that they have no competing interests.

## Supplementary Figures

**Supplementary Figure 1.**
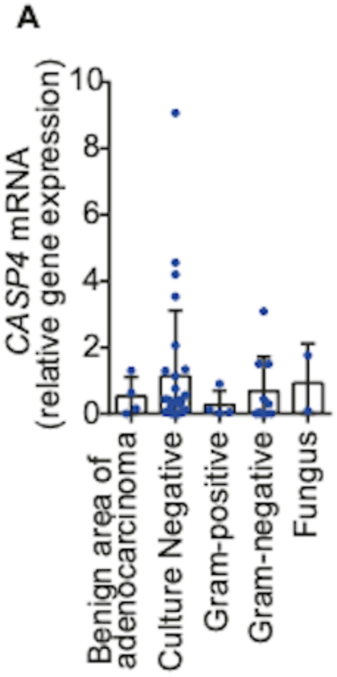
Caspase-4 is expressed in lung autopsy of lethal COVID19 patients regardless of bacterial and fungal co-infections. Lung autopsies were obtained from fatal cases of COVID-19 were distributed according to presence of absence of co-infections as determined by blood culture during hospitalization. COVID19 patients includes 28 that tested negative for co-infections, 4 coinfected with Gram-positive bacteria, 9 coinfected with Gram-negative bacteria and 2 coinfected with fungus. The data showing 28 that tested negative for co-infections and 4 patients controls that deceased due to lung adenocarcinoma (benign area of lungs) were already presented in Fig. 2A. The expression of caspase-4 mRNA (*CASP4*) was assessed by RT-PCR. Box shows average ± SD of the values.

**Supplementary Figure 2.**
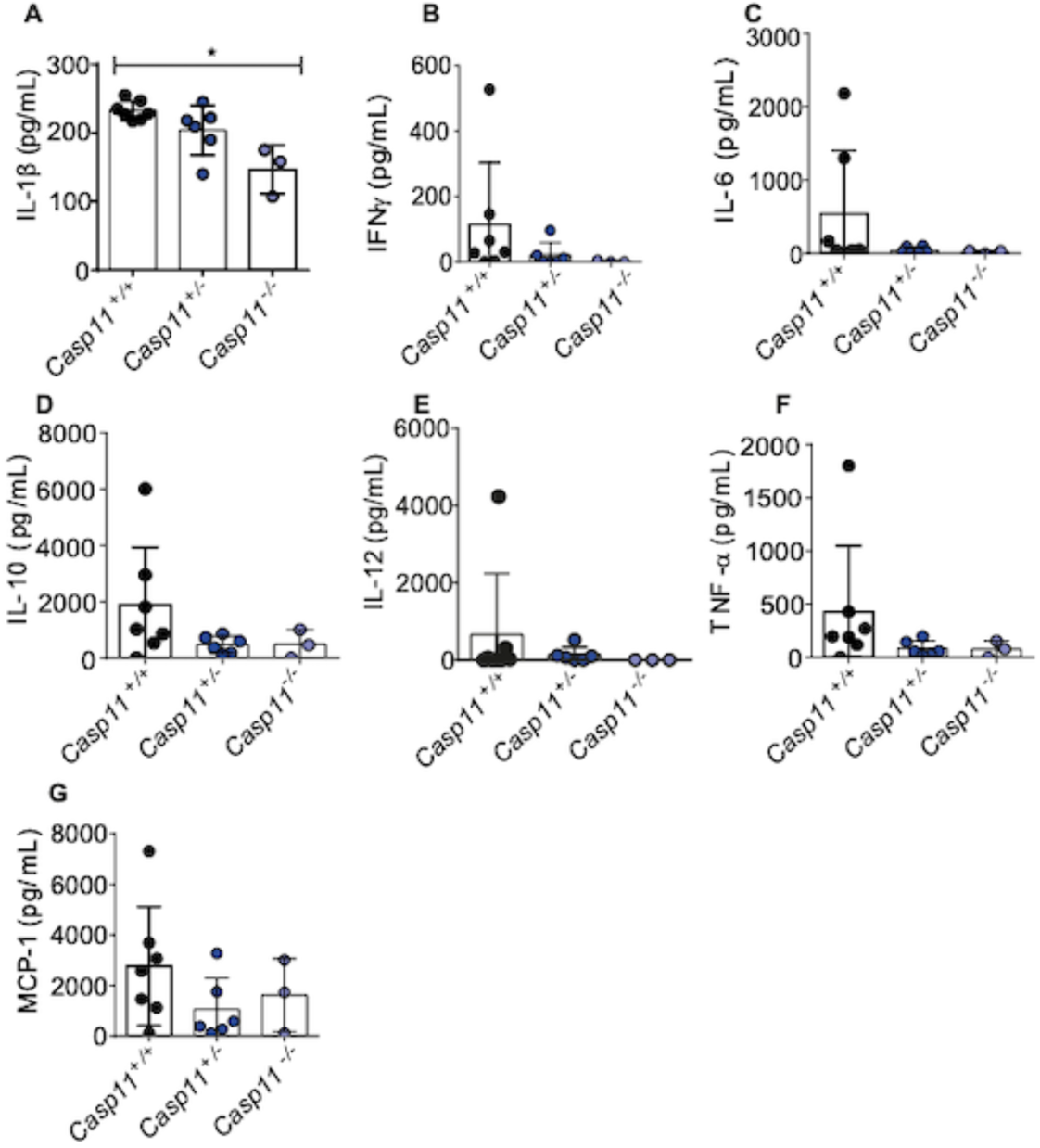
Cytokine production in the lungs of mice after infection with SARS-CoV-2. Transgenic K18-hACE2 mice heterozygous for *Casp11* were crossed to generate K18-hACE2 *Casp11^+/+^*, *Casp11^+/–^* and *Casp11^−/−^* littermate control mice. Animals were infected intranasally with 2×10^4^ SARS-CoV-2 per mice for 3 days and the cytokines IL-1β (A), IFNγ (B), IL-6 (C), IL-10 (D), IL-12 (E), TNFα (F), and MCP-1 (G) were evaluated in lung tissue homogenate by CBA Mouse Inflammatory Cytokine Kit (BD) following instructions. * *P* < 0.05, as determined Kruskal-Wallis followed by Dunn post hoc test. Box shows average ± SD of the values.

**Supplementary Figure 3.**
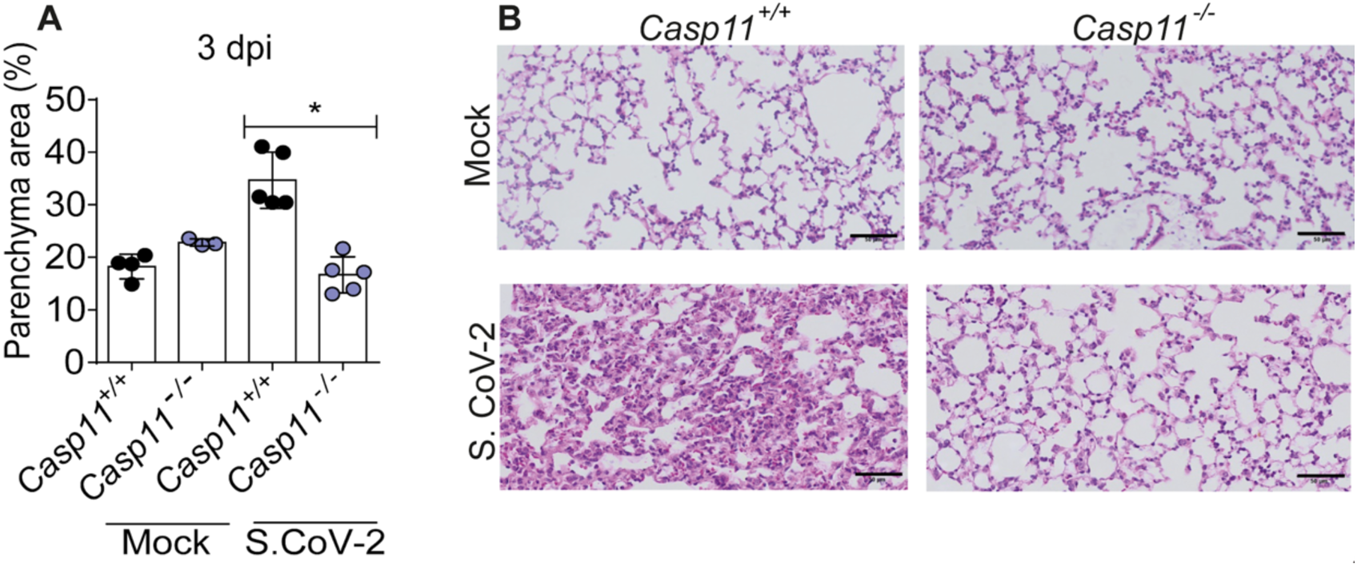
Caspase-11 aggravates disease pathology, assessed by histological analyzes of lungs from mice infected with SARS-CoV-2. Transgenic K18-hACE2 mice heterozygous for *Casp11* were crossed to generate K18-hACE2 *Casp11^−/−^* mice and littermate controls. Animals were infected intranasally with 2×10^4^ SARS-CoV-2 per mice for 3 days. (A) The percentage of parenchyma area were estimated after hematoxylin and eosin (H&E) staining of lung sections of K18-hACE2 *Casp11^+/+^*, and *Casp11^−/−^* infected for 3 days. Box shows average ± SD of the values. Representative images of H&E-stained lungs of mice infected for 3 days are shown (B). Upper panels show Mock-infected mice. Scale bar 50 μm. *, *P* < 0.05, as determined by Man Whitney test.

**Supplementary Table 1.** Clinical Score evaluated in infected mouse

| **Variable** | **Clinical correlate** | **Score** |
| --- | --- | --- |
| Weight | 0% of loss | 0 |
|  | 1 to 5% of loss | 1 |
|  | 6 to 10% of loss | 2 |
|  | 11 to 15% of loss | 3 |
|  | 16 to 20% of loss | 4 |
|  | >20%of loss | 5 |
| Posture and appearance | Without piloerection | 0 |
|  | With piloerection | 1 |
| Activity | Active | 0 |
|  | Moderate activity | 1 |
|  | Few activity | 2 |
|  | Immobile | 3 |
| Eyes opening | Normal | 0 |
|  | Slow closing | 1 |
|  | Closed | 2 |
| Responsiveness to stimuli | Normal | 0 |
|  | Moderate | 1 |
|  | Low | 2 |
| Breath | Normal | 0 |
|  | Difficulty breathing | 1 |

## Notes

### Competing Interest Statement

The authors have declared no competing interest.

### Funding Statement

Fundacao de Amparo a Pesquisa do Estado de Sao Paulo, FAPESP grants 2013/08216-2, 2019/11342-6 and 2020/04964-8. Conselho Nacional de Desenvolvimento Cientifico e Tecnologico, CNPq grant 303021/2020-9. Coordenacao de Aperfeicoamento de Pessoal de Nivel Superior, CAPES grant 88887.507253/2020-00.

### Author Declarations

Research Ethics Committee Protocol from Hospital das Clinicas de Ribeirao Preto USP gave ethical approval for this work.

